# Distinguishing COVID-19 from influenza pneumonia in the early stage through CT imaging and clinical features

**DOI:** 10.1101/2020.04.17.20061242

**Authors:** Zhiqi Yang, Daiying Lin, Xiaofeng Chen, Jinming Qiu, Shengkai Li, Ruibin Huang, Hongfu Sun, Yuting Liao, Jianning Xiao, Yanyan Tang, Guorui Liu, Renhua Wu, Xiangguang Chen, Zhuozhi Dai

**Affiliations:** Department of Radiology, Meizhou People’s Hospital, Guangdong, 514031, P. R. China; Department of Radiology, Shantou Central Hospital, Shantou, Guangdong, 515041, P. R. China; Department of Radiology, Second Affiliated Hospital, Shantou University Medical College, Shantou, Guangdong, 515000, P. R. China; Department of Radiology, Huizhou Municipal Central Hospital, Huizhou, Guangdong 516001, China; Department of Radiology, First Affiliated Hospital, Shantou University Medical College, Shantou, Guangdong, 515041, P. R. China; School of Information Technology and Electrical Engineering, University of Queensland, Queensland, 4072, Australia; GE Healthcare, Guangzhou 510623, China

**Keywords:** COVID-19, Influenza, Radiology, Diagnosis

## Abstract

**Purpose:** To identify differences in CT imaging and clinical features between COVID-19 and influenza pneumonia in the early stage, and to identify the most valuable features in the differential diagnosis.

**Materials and Method:** A consecutive cohort of 73 COVID-19 and 48 influenza pneumonia patients were retrospectively recruited from five independent institutions. The courses of both diseases were confirmed to be in the early stages (2.66 ± 2.62 days for COVID-19 and 2.19 ± 2.10 days for influenza pneumonia after onset). The chi-square test, student’s *t*-test, and Kruskal-Wallis *H*-test were performed to compare CT imaging and clinical features between the two groups. Spearman or Kendall correlation tests between feature metrics and diagnosis outcomes were also assessed. The diagnostic performance of each feature in differentiating COVID-19 from influenza pneumonia was evaluated with univariate analysis. The corresponding area under the curve (AUC), accuracy, specificity, sensitivity and threshold were reported.

**Results:** The ground-glass opacification (GGO) was the most common imaging feature in COVID-19, including pure-GGO (75.3%) and mixed-GGO (78.1%), mainly in peripheral distribution. For clinical features, most COVID-19 patients presented normal white blood cell (WBC) count (89.04%) and neutrophil count (84.93%). Twenty imaging features and 6 clinical features were identified to be significantly different between the two diseases. The diagnosis outcomes correlated significantly with the WBC count (r=-0.526, *P*<0.001) and neutrophil count (r=-0.500, *P*<0.001). Four CT imaging features had absolute correlations coefficients higher than 0.300 (*P*<0.001), including crazy-paving pattern, mixed-GGO in peripheral area, pleural effusions, and consolidation.

**Conclusions:** Among a total of 1537 lesions and 62 imaging and clinical features, 26 features were demonstrated to be significantly different between COVID-19 and influenza pneumonia. The crazy-paving pattern was recognized as the most powerful imaging feature for the differential diagnosis in the early stage, while WBC count yielded the highest diagnostic efficacy in clinical manifestations.

## Introduction

The coronavirus disease 2019 (COVID-19) pandemic is a global crisis, which has killed more than seventy thousand peoples as of April 7, 2020 (1). A clear picture of imaging and clinical manifestations of COVID-19 remains unknown. These manifestations of COVID-19 are protean and usually overlap with those of other viral pneumonia (2, 3). In the early stage of COVID-19, the main radiological finding is the ground-glass opacity (GGO), especially the pure ground-glass opacity (2), in the subpleural region, located unilaterally or bilaterally in the lower lobes (3). The lesions can develop one or more lobes, with a slight preference for the lower right lobe (4). However, these CT imaging findings are similar to those of influenza pneumonia (5, 6). The main clinical manifestations of COVID-19, including fever, dry cough, and fatigue, are also non-specific (7, 8).

Both COVID-19 and influenza pneumonia are highly contagious and present similar symptoms. The US CDC has reported that some COVID-19 deaths have been miscategorized as influenza (9). Unlike for influenza, no vaccine or antiviral agents are available for COVID-19 at the moment (10). Moreover, the mortality rate for COVID-19 appears to be substantially higher than for influenza, about 5.6% vs 0.1% based on the primary data (1). Therefore, the discrimination between COVID-19 and influenza is critical in clinical practice. Accurate imaging and clinical feature recognition can aid in early diagnosis for COVID-19 and thus prevent spreading and speed up treatment.

In our previous study, we demonstrated that based on CT imaging and clinical manifestations alone, the pneumonia patients with and without COVID-19 can be distinguished (11). Harrison et al. examined the performance of seven radiologists in differentiating COVID-19 from viral pneumonia on chest CT results and found the average sensitivity of 80% and specificity of 84% (12). However, we realized that about 44% of the viral pneumonia cases were Human Rhinovirus, and influenza pneumonia only accounted for about 15%. To our knowledge, no study has explored the differences between COVID-19 and influenza using CT imaging and clinical features. In this study, we aim to identify differences in CT imaging and clinical features between COVID-19 and influenza pneumonia in the early stage, and to identify the most valuable features in distinguishing COVID-19 from influenza pneumonia, based on multi-center data.

## Materials and Methods

### Patients

Ethical approval by the institutional review boards of Second affiliated Hospital of Shantou University Medical College (Approval number: SDYFE202029) was obtained for this retrospective analysis, with the requirement for informed consent waived. From January 1 to February 15 in the year 2020, 73 consecutive patients confirmed with severe acute respiratory syndrome coronavirus 2 infection by real-time reverse transcription polymerase chain reaction (RT-PCR) from 5 independent hospitals in 4 Chinese cities were enrolled in this study. Of all the patients, including 24 from Huizhou city, 25 from Shantou city, 15 from Yongzhou city and 9 from Meizhou city, the mean age was 41.9 years (range: 3 - 69 years). Among them, 41 patients were men (mean age: 41.4 years; range: 16 - 69 years) and 32 were women (mean age: 42.6 years; range: 3 - 66 years).

In addition, from January 1 2015 to September 30 2019, a total of 205 consecutive patients confirmed with influenza pneumonia from Shantou and Meizhou city were recruited. **Figure 1** showed the patient recruitment pathway for the influenza pneumonia group, along with the inclusion and exclusion criteria. According to the 2019-2020 guide from the Chinese center for disease control and prevention (13), 137 patients were confirmed to have influenza A virus infection, 68 patients with influenza B virus infection. 101 patients with influenza A virus infection and 56 patients with influenza B virus infection were excluded because of they did not have chest CT or clinical data. Finally, 48 influenza pneumonia patients (mean age: 40.4 years, range: 0.1 - 83 year) were enrolled as controls, including 30 men (mean age: 40.1 years; range: 0.1 - 72 years) and 18 women (mean age:40.8 years; range: 0.1 - 83 years). Among them, 12 patients with influenza B virus infection (mean age: 19.0 years, range: 0.1 - 63 years) and 36 patients with influenza A virus infection (mean age: 47.5 years, range: 0.2 - 83 years).

**Figure 1.**
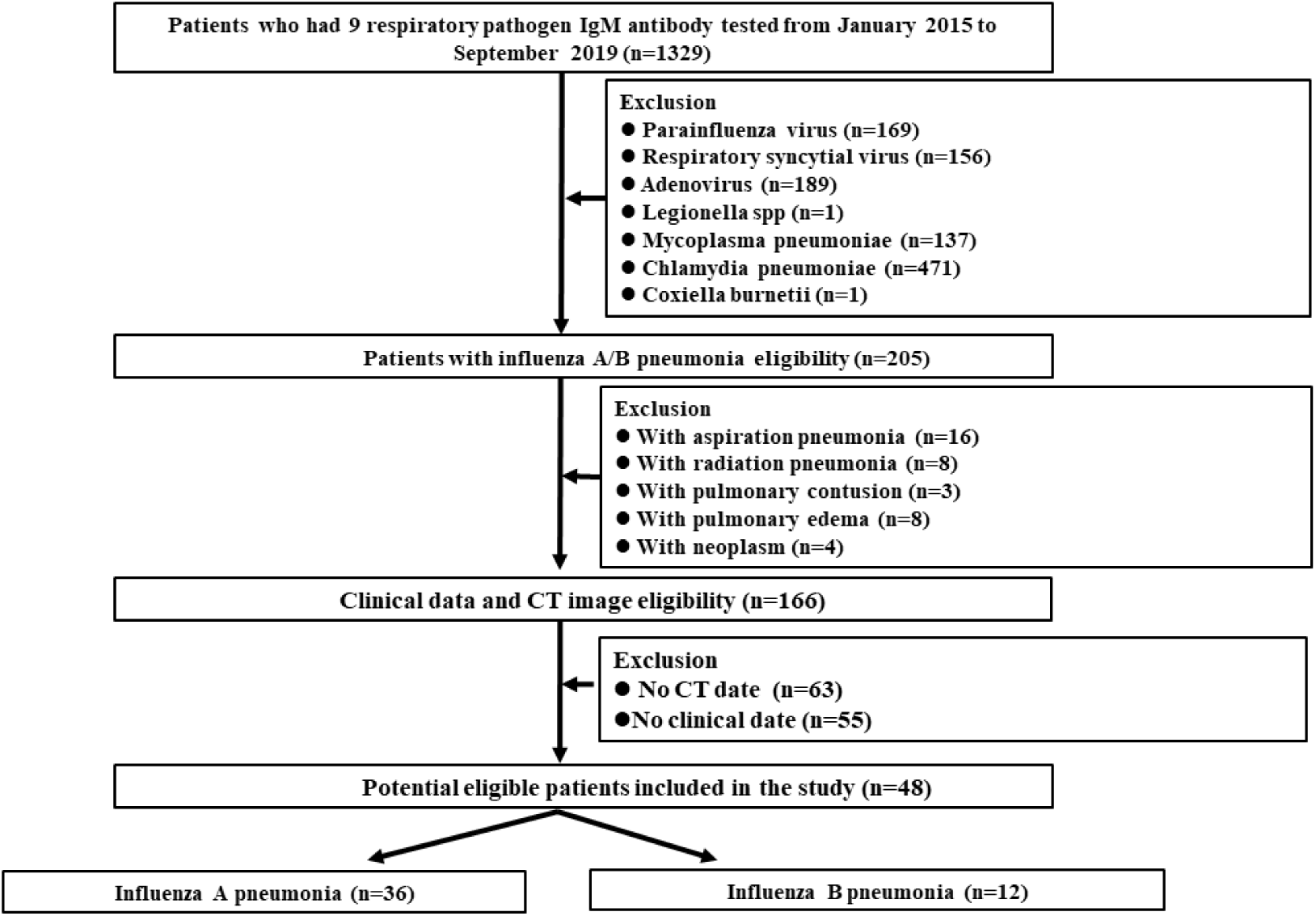
Flowchart shows influenza pneumonia patient selection, along with the inclusion and exclusion criteria.

### Image and clinical data collection

Non-contrast-enhanced chest CT imaging data were obtained from multiple hospitals of varied CT systems, including GE CT Discovery 750 HD (General Electric, US), SCENARIA 64 CT (Hitachi Medical, Japan), PHILIPS Ingenuity CT (PHILIPS, Netherlands), and Siemens SOMATOM Definition AS (Siemens, Germany) systems. All images were reconstructed into 1 mm slices with a slice interval of 0.8 mm. The detailed acquisition parameters are summarized in the supplementary material (**Table E1**).

The baseline clinical data including course of disease, age, gender, body temperature, clinical symptoms (including cough, fatigue, sore throat, stuffy, and runny nose), total white blood cell (WBC) count, lymphocyte count, lymphocyte ratio, neutrophil count, neutrophil ratio and c-reactive protein (CRP) level were collected. According to the normal range used at individual hospital, the threshold value for WBC count, lymphocyte count, lymphocyte ratio, neutrophil count, neutrophil ratio and CRP level was set to 3.5∼9.5×10^9^/L, 1.1∼3.2×10^9^/L, 20.0∼50.0%,1.8∼6.3 ×10^9^/L,40.0∼75.0% and 0.0∼6.0 mg/L, respectively.

### CT Image analysis

A total of 26 quantitative and 22 qualitative imaging features were extracted for analysis. The descriptions of the CT imaging features are listed in the supplementary material (**Table E2**). For the extraction of CT qualitative and quantitative imaging features, two senior radiologists (Z.Y. and X.C., more than 15 years of experience) reached a consensus and were blinded to the clinical and laboratory findings. Lesion in the outer third of the lung was defined as peripheral and lesion in the inner two thirds of the lung was defined as central. The classification of the lesion size is based on a previous study (14). The progression of lesion within each lung lobe was evaluated by scoring each lobe from 0 to 4 (15), corresponding to normal, 1% ∼25% infection, 26%∼ 50% infection, 51%∼ 75% infection and more than 75% infection, respectively. The scores were combined for all 5 lobes to provide a total score ranging from 0 to 20. **Figure 2** was one example of the evaluation of chest CT images.

**Figure 2.**
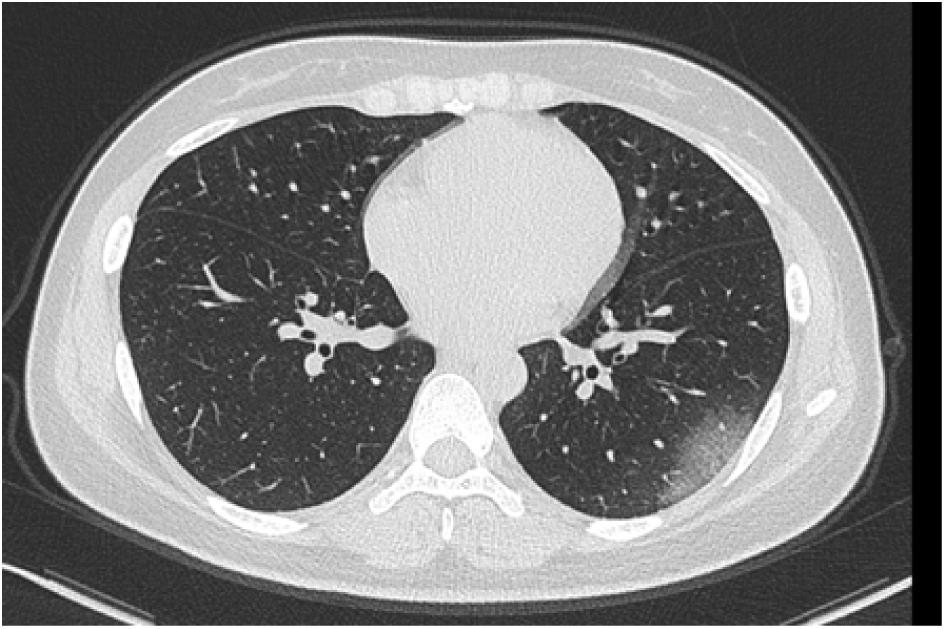
Axial non contrast-enhanced CT image from a 26-year old female patient with COVID-19. Pure ground glass opacities are observed in peripheral area in left lower lobe. The maximum diameter of lesion is 4.5 cm. The left lower lobe score is 1 because of the involve lung parenchyma less than 25%.

### Statistical Analysis

The CT imaging and clinical features were compared between COVID-19 and influenza pneumonia group by using the chi-square test (for nominal variable), the Kruskal-Wallis *H* test (for ordinal variable), or the student’s *t* test (for continuous variable). The features with a significant difference between the two groups were extracted. Spearman or Kendall correlation test between feature metrics and diagnosis outcomes (i.e., 1 for COVID-19 and 0 for influenza pneumonia) were assessed for each extracted feature. The diagnostic performance of clinical and CT features in differentiating COVID-19 from influenza pneumonia was evaluated with univariate analysis. Additionally, corresponding area under the curve (AUC), accuracy, specificity, sensitivity and threshold were calculated. All statistical analyses for this study were performed with R (version 3.6.4, http://www.r-project.org/). A two-tailed *P*-value < 0.05 indicated statistical significance.

## Results

### Clinical features comparison between groups

121 patients, including 73 COVID-19 and 48 influenza pneumonia were recruited in this study. The courses of both diseases were confirmed to be in the early stages, which were 2.66 ± 2.62 days for COVID-19 and 2.19 ± 2.10 days for influenza pneumonia after onset. A total of 15 clinical features of COVID-19 and influenza pneumonia patients are shown in **Table 1**. Compare to COVID-19 patients, influenza pneumonia patients have higher temperature (*P* < 0.001), WBC count (*P* < 0.001), neutrophil count (*P* < 0.001), neutrophil rate (*P*=0.017), CRP level (*P*=0.033) and have lower lymphocyte rate (*P*=0.005). There is no significant difference in sex, age, cough, fatigue, sore throat, stuffy, runny nose, and lymphocyte count between the two groups. As shown in Figure 3, most COVID-19 patients present normal WBC count (89.04%), neutrophil count (84.93%) and neutrophil rate (63.01%).

**Table 1:**
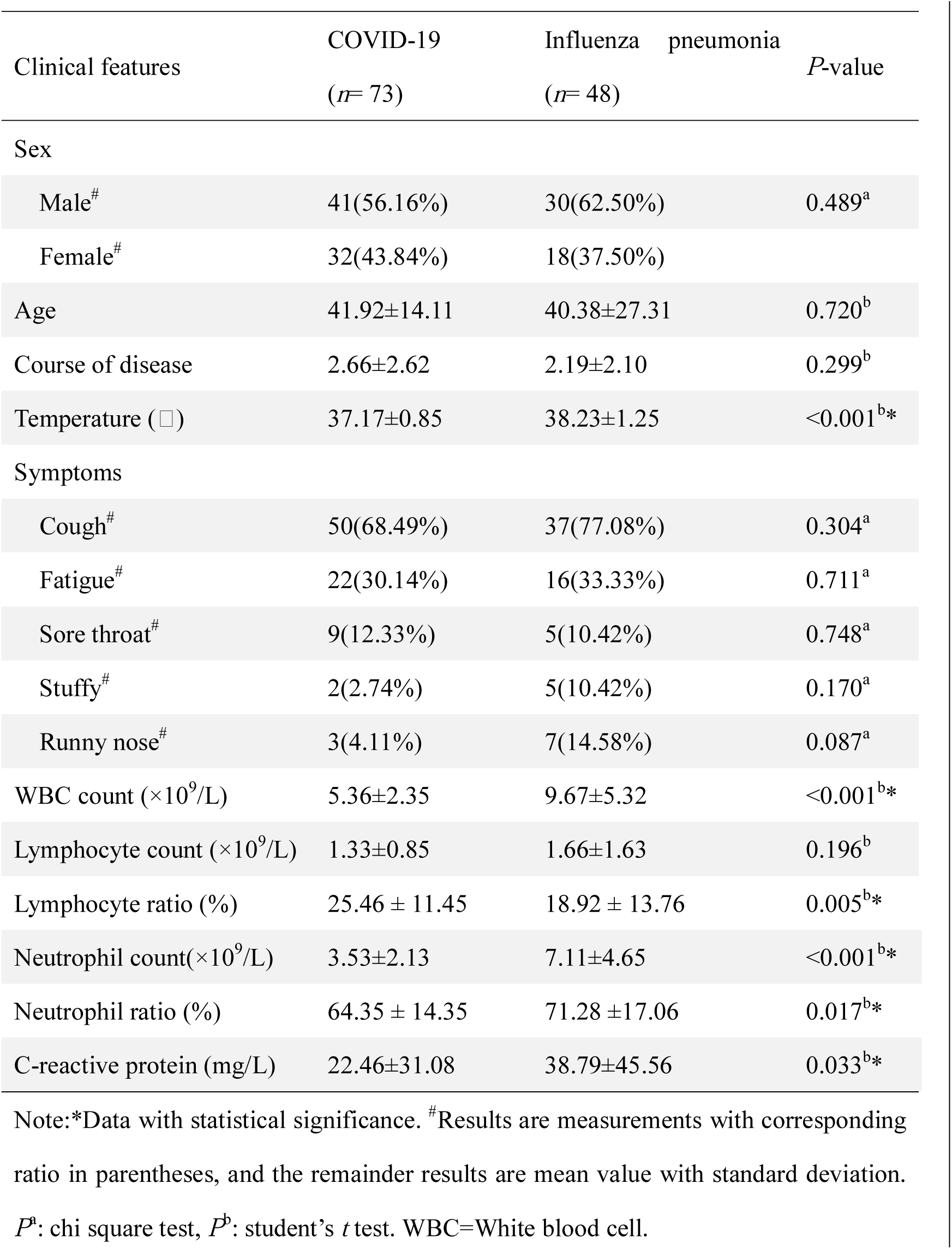
Clinical features of COVID-19 and influenza pneumonia patients

| Clinical features | COVID-19<br>(n= 73) | Influenza pneumonia<br>(n= 48) | P-value |
| --- | --- | --- | --- |
| Sex |  |  |  |
| Male<sup>#</sup> | 41(56.16%) | 30(62.50%) | 0.489<sup>a</sup> |
| Female<sup>#</sup> | 32(43.84%) | 18(37.50%) |  |
| Age | 41.92±14.11 | 40.38±27.31 | 0.720<sup>b</sup> |
| Course of disease | 2.66±2.62 | 2.19±2.10 | 0.299<sup>b</sup> |
| Temperature (□) | 37.17±0.85 | 38.23±1.25 | <0.001<sup>b*</sup> |
| Symptoms |  |  |  |
| Cough<sup>#</sup> | 50(68.49%) | 37(77.08%) | 0.304<sup>a</sup> |
| Fatigue<sup>#</sup> | 22(30.14%) | 16(33.33%) | 0.711<sup>a</sup> |
| Sore throat<sup>#</sup> | 9(12.33%) | 5(10.42%) | 0.748<sup>a</sup> |
| Stuffy<sup>#</sup> | 2(2.74%) | 5(10.42%) | 0.170<sup>a</sup> |
| Runny nose<sup>#</sup> | 3(4.11%) | 7(14.58%) | 0.087<sup>a</sup> |
| WBC count (×10<sup>9</sup>/L) | 5.36±2.35 | 9.67±5.32 | <0.001<sup>b*</sup> |
| Lymphocyte count (×10<sup>9</sup>/L) | 1.33±0.85 | 1.66±1.63 | 0.196<sup>b</sup> |
| Lymphocyte ratio (%) | 25.46 ± 11.45 | 18.92 ± 13.76 | 0.005<sup>b*</sup> |
| Neutrophil count(×10<sup>9</sup>/L) | 3.53±2.13 | 7.11±4.65 | <0.001<sup>b*</sup> |
| Neutrophil ratio (%) | 64.35 ± 14.35 | 71.28 ±17.06 | 0.017<sup>b*</sup> |
| C-reactive protein (mg/L) | 22.46±31.08 | 38.79±45.56 | 0.033<sup>b*</sup> |

Note: \*Data with statistical significance. <sup>#</sup>Results are measurements with corresponding ratio in parentheses, and the remainder results are mean value with standard deviation.
P<sup>a</sup>: chi square test, P<sup>b</sup>: student's *t* test. WBC=White blood cell.

**Figure 3.**
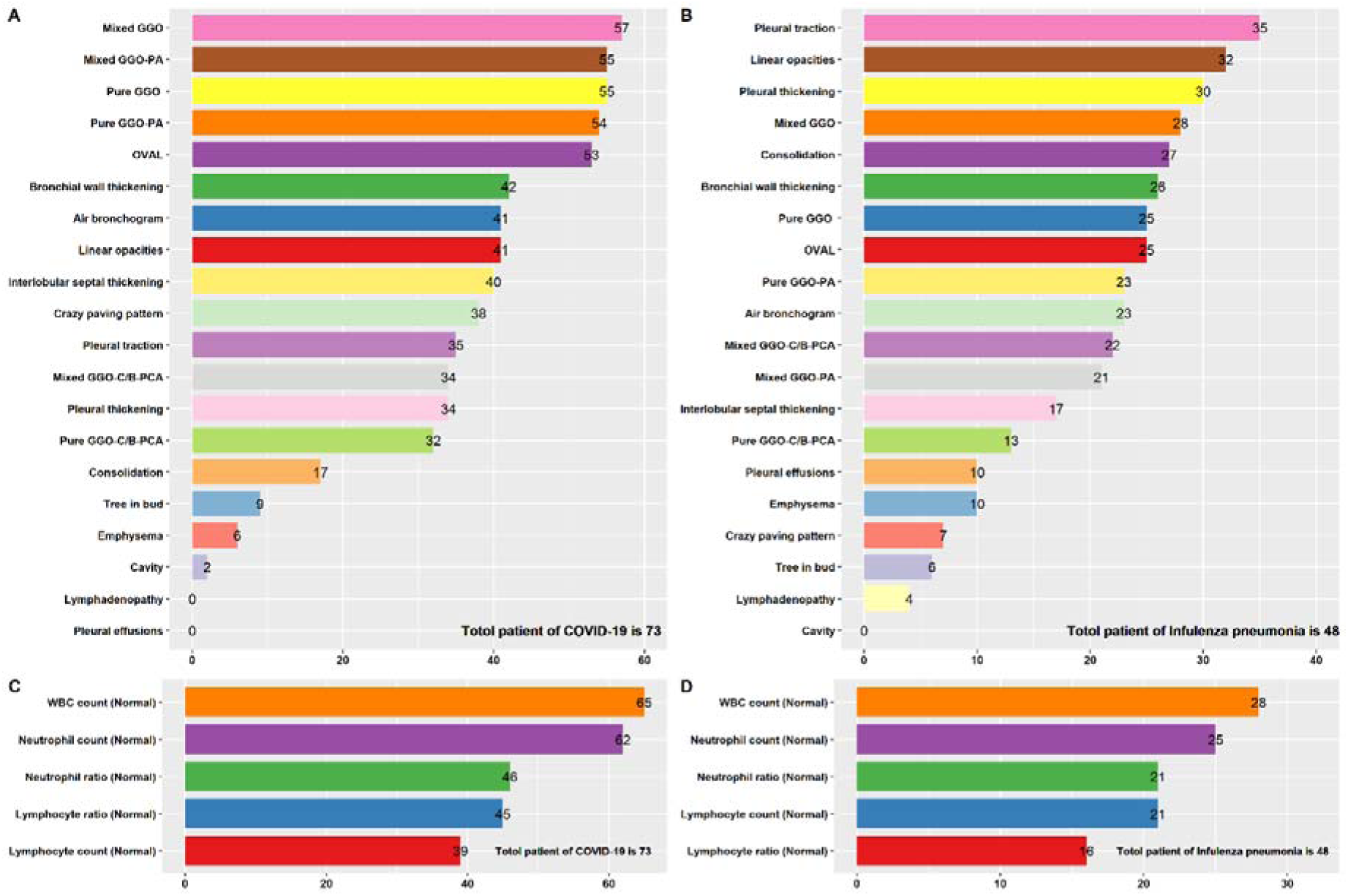
CT and clinical features distribution of COVID-19 and influenza patients. A: CT features of distribution of COVID-19 patients; B: CT features of distribution of influenza patients; C: Clinical features distribution of COVID-19 patients, and D: Clinical features distribution of influenza patients. The number of patients with corresponding feature was presented on the right side of horizontal axis. WBC = white blood cell, GGO = Ground-glass opacification, Mixd GGO-PA = Mixed GGO in peripheral area, OVAL = Offending vessel augmentation in lesions, Mixd GGO-C/B-PAC = Mixd GGO in central/both peripheral and central area, Pure GGO-C/B-PAC = Pure GGO in central / both peripheral and central area.

### Imaging features comparison between groups

A total of 1537 lesions were identified, with 1073 from COVID-19 group and 464 from influenza pneumonia group. The differences between COVID-19 and influenza pneumonia patients for CT quantitative and qualitative imaging features were showed in supplementary materials **Table E3 and Table E4**, respectively. Those features with significant differences were presented in Table 2. For imaging manifestations, 9 patients in the COVID-19 group (12.33%) and 3 patients in the influenza pneumonia group (6.25%) showed normal chest CT. Of all quantitative imaging features, COVID-19 patients have a greater total number of pure GGO (*P* = 0.01), total number of pure GGO in peripheral area (*P* = 0.003), total number of mixed GGO in peripheral area (*P*=0.016), and total number of lesions in peripheral area (*P* = 0.003). However, COVID-19 patients have a fewer total number of consolidation (*P* = 0.018) and total scores of left lung (*P* = 0.032). Compared to influenza pneumonia patients, more lesions are between 1 cm to 3 cm (*P* = 0.005) in COVID-19 patients.

**Table 2:** CT imaging features with significant differences between COVID-19 and influenza pneumonia patients

| Imaging features | COVID-19<br>(n=73) | influenza<br>pneumonia(n=48) | P-value |
| --- | --- | --- | --- |
| Number of pure GGO |  |  |  |
| Total | 6.78±11.28 | 2.75±5.33 | 0.010<sup>b*</sup> |
| Peripheral area | 4.81±7.15 | 1.92±3.16 | 0.003<sup>b*</sup> |
| Number of mixed GGO in peripheral area | 4.60±6.92 | 2.15±4.12 | 0.016<sup>b*</sup> |
| Number of consolidations | 0.60±1.65 | 1.60±2.52 | 0.018<sup>b*</sup> |
| Total number of lesions in peripheral area | 10.74±13.69 | 5.15±6.63 | 0.003<sup>b*</sup> |
| Lesion sizes (1cm to 3cm) | 8.29±14.24 | 3.21±4.19 | 0.005<sup>b*</sup> |
| Total scores of involved lung zones |  |  |  |
| Left lung | 2.15±1.86 | 3.10±2.62 | 0.032<sup>b*</sup> |
| Bilateral lower lobes | 2.59±2.18 | 3.69±2.52 | 0.015<sup>b*</sup> |
| Pure GGO<sup>#</sup> |  |  | 0.008<sup>a*</sup> |
| Negative | 18(24.7%) | 23(47.9%) |  |
| Positive | 55(75.3%) | 25(25.1%) |  |
| Pure GGO in peripheral area<sup>#</sup> |  |  | 0.004<sup>a*</sup> |
| Negative | 19(26.0%) | 25(52.1%) |  |
| Positive | 54(74.0%) | 23(47.9%) |  |
| Mixed GGO<sup>#</sup> |  |  | 0.020<sup>a*</sup> |
| Negative | 16(21.9%) | 20(41.7%) |  |
| Positive | 57(78.1%) | 28(58.3%) |  |
| Mixed GGO in peripheral area<sup>#</sup> |  |  | <0.001<sup>a*</sup> |
| Negative | 18(24.7%) | 27(56.3%) |  |
| Positive | 55(75.3%) | 21(43.7%) |  |
| Consolidation<sup>#</sup> |  |  | <0.001<sup>a*</sup> |
| Negative | 56(76.7%) | 21(43.8%) |  |
| Positive | 17(23.3%) | 27(56.2%) |  |
| Interlobular septal thickening<sup>#</sup> |  |  | 0.037<sup>a*</sup> |
| Negative | 33(45.21%) | 31(64.58%) |  |
| Positive | 40(54.79%) | 17(35.42%) |  |
| Crazy paving pattern<sup>#</sup> |  |  | <0.001<sup>a*</sup> |
| Negative | 35(47.95%) | 41(85.42%) |  |
| Positive | 38(52.05%) | 7(14.58%) |  |
| Offending vessel augmentation in lesions<sup>#</sup> |  |  | 0.021<sup>a*</sup> |
| Negative | 20(27.40%) | 23(47.92%) |  |
| Positive | 53(72.60%) | 25(52.08%) |  |
| Pleural traction<sup>#</sup> |  |  | 0.007<sup>a*</sup> |
| Negative | 38(52.05%) | 13(27.08%) |  |
| Positive | 35(47.95%) | 35(72.92%) |  |
| Emphysema<sup>#</sup> |  |  | 0.045<sup>a*</sup> |
| Negative | 67(91.78%) | 38(79.17%) |  |
| Positive | 6(8.22%) | 10(20.83%) |  |
| Pleural effusions<sup>#</sup> |  |  | <0.001<sup>a*</sup> |
| Negative | 73(100.00%) | 38(79.17%) |  |
| Positive | 0(0.00%) | 10(20.83%) |  |
| Lymphadenopathy<sup>#</sup> |  |  | 0.047<sup>a*</sup> |
| Negative | 73(100.00%) | 44(91.67%) |  |
| Positive | 0(0.00%) | 4(8.33%) |  |

Note: \* Data with statistical significance. <sup>#</sup> Results are measurements with corresponding ratio in parentheses, and the remainder results are mean value with standard deviation.
*P*<sup>a</sup>: chi square test. *P*<sup>b</sup>: student's *t* test. GGO = ground-glass opacification

For all qualitative imaging features, most COVID patients present higher positive rate of interlobular septal thickening (54.79%), crazy paving pattern (52.05%), offending vessel augmentation in lesions (72.60%) and lower positive rate of pleural traction (47.95%), emphysema (8.22%), pleural effusions (0.00%), lymphadenopathy (0.00%). The ranking of these features was shown in **Figure 3**. Compared to the COVID-19 patients, the decreased positive rate of interlobular septal thickening (35.42%), crazy paving pattern (14.58%), offending vessel augmentation in lesions (52.08%), as well as increased positive rate of pleural traction (72.92%), emphysema(20.83%), pleural effusions (20.83%), lymphadenopathy (8.83%) are more pronounced in influenza virus infection patients (all *P*< 0.05).

### Correlation analysis and diagnostic performance

The correlation analysis and diagnostic performance of clinical features in distinguishing COVID-19 from influenza pneumonia were shown in **Table 3**. The diagnosis outcomes correlated significantly with the WBC count (Spearman’s r correlation, r = −0.526, P < 0.001) and neutrophil count (r = −0.500, P < 0.001). Lymphocyte rate and temperature have a weaker correlation with distinguishing COVID-19 from influenza pneumonia, with r = 0.310 (P < 0.001) and r = −0.433 (P < 0.001), respectively. However, little correlations were found for C-reactive protein and for neutrophil ratio in differential diagnosis. The WBC count yield a maximum AUC of 0.811 (95% CI: 0.731 ∼ 0.890), follow by neutrophil count with the AUC of 0.795 (95% CI: 0.711 ∼ 0.879). The distribution of WBC count and neutrophil count in both groups were shown in **Figure 4**.

**Table 3:** Correlation analysis and diagnostic performance of clinical features in distinguishing COVID-19from influenza pneumonia

| Clinical features | Correlation analysis |  | ROC analysis |  |  |  |  |  |
| --- | --- | --- | --- | --- | --- | --- | --- | --- |
|  | r | P-value | AUC | 95% CI | Accuracy | Specificity | Sensitivity | Threshold |
| Lymphocyte ratio | 0.310<sup>a</sup> | <0.001 | 0.683 | 0.581~0.785 | 0.686 | 0.616 | 0.792 | 23.65 |
| C-reactive protein | -0.204<sup>a</sup> | 0.025 | 0.620 | 0.517~0.724 | 0.661 | 0.822 | 0.417 | 34.82 |
| Neutrophil ratio | -0.264<sup>a</sup> | 0.003 | 0.656 | 0.552~0.760 | 0.669 | 0.658 | 0.688 | 65.78 |
| Temperature | -0.433<sup>a</sup> | <0.001 | 0.755 | 0.663~0.847 | 0.744 | 0.890 | 0.521 | 38.15 |
| Neutrophil count | -0.500<sup>a</sup> | <0.001 | 0.795 | 0.711~0.879 | 0.769 | 0.822 | 0.688 | 4.610 |
| WBC count | -0.526<sup>a</sup> | <0.001 | 0.811 | 0.731~0.890 | 0.760 | 0.781 | 0.729 | 6.435 |
| Note: <sup>a</sup> r and corresponding P-value are computed by Spearman's correlation test.CI = Confidence Interval. WBC = White blood cell. |  |  |  |  |  |  |  |  |

**Figure 4.**
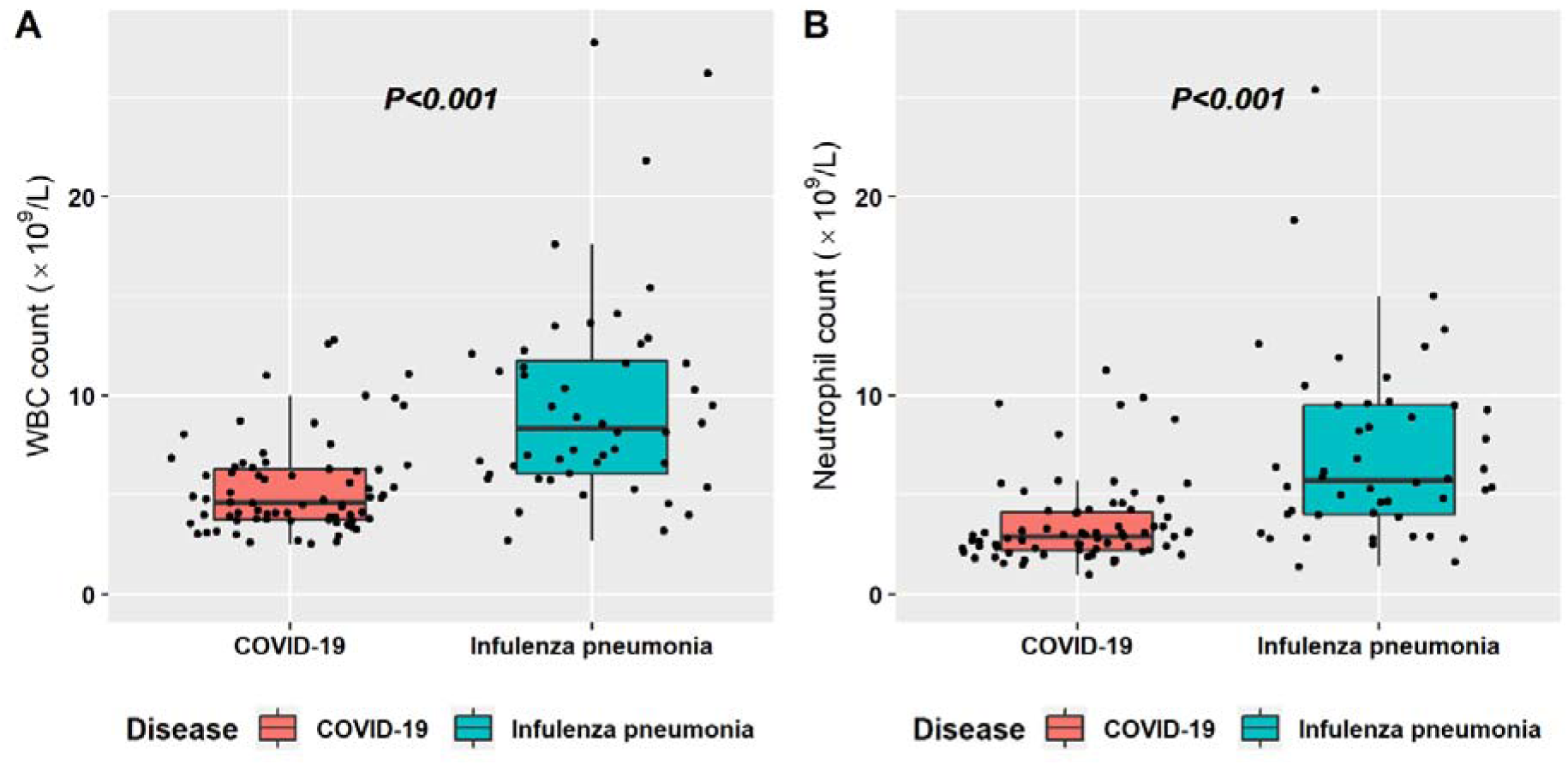
Box plot graphs revealing statistically significant differences both in the white blood cell (WBC) count (A) and in neutrophil count (B) between COVID-19 and influenza pneumonia patients. Most cases in both diseases have normal WBC count and neutrophil count, however, the overall values in influenza pneumonia are higher than those in COVID-19 (*P* < 0.001).

The correlation analysis and diagnostic performance of CT features in distinguishing COVID-19 from influenza pneumonia were shown in Table 4. In COVID-19 diagnosis, the crazy paving pattern achieved the highest correlation of 0.379 (P < 0.001), which had an AUC of 0.687 (95% CI: 0.611 ∼ 0.764). Mixed GGO in peripheral area had a correlation of 0.320 (P < 0.001). The consolidation and pleural effusions were more common in influenza pneumonia compared to COVID-19. The correlations for consolidation and for pleural effusions were −0.335 (P < 0.001) and −0.370 (P < 0.001), respectively. The typical CT imaging features of both diseases were illustrated in **Figure 5**.

**Table 4:**
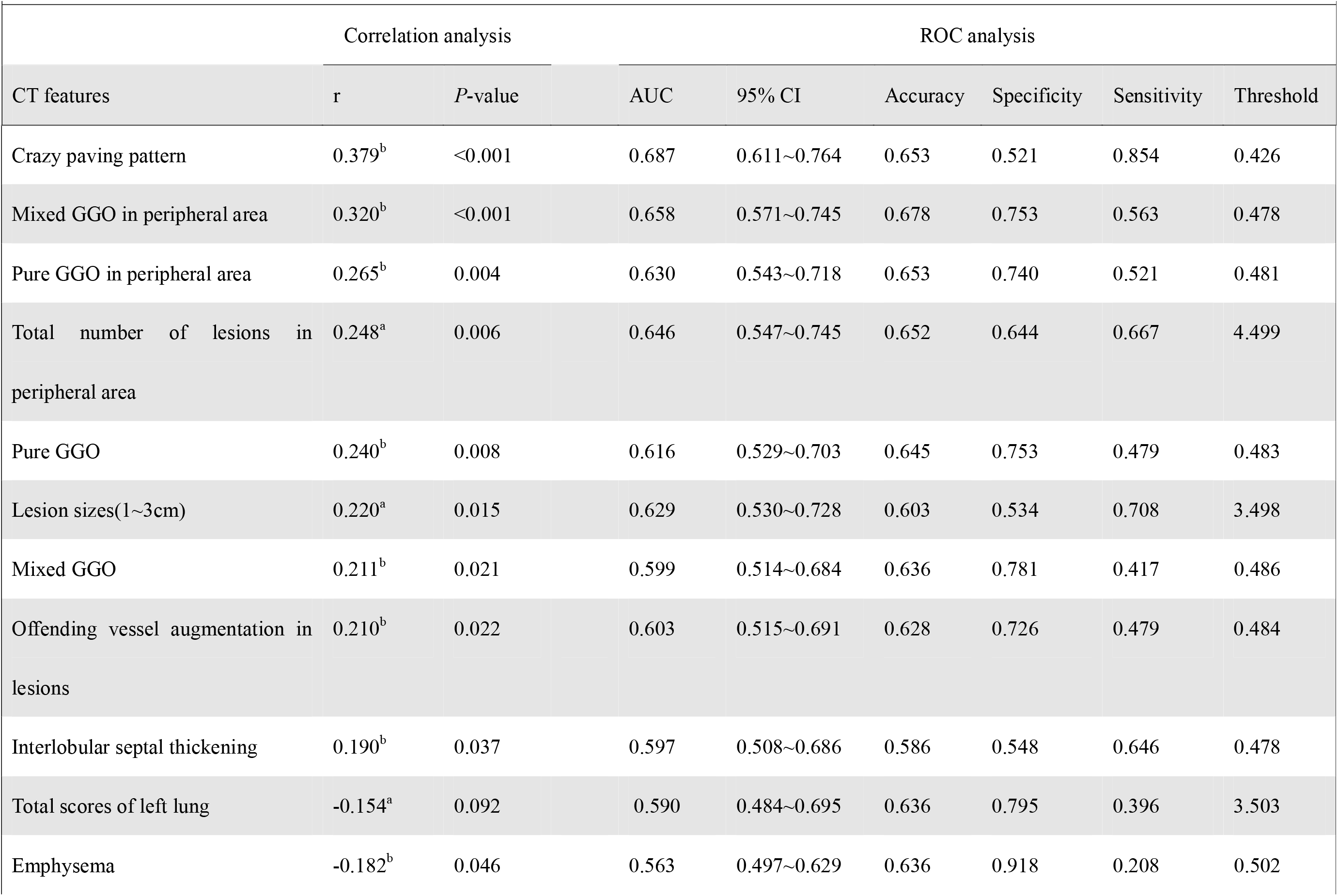
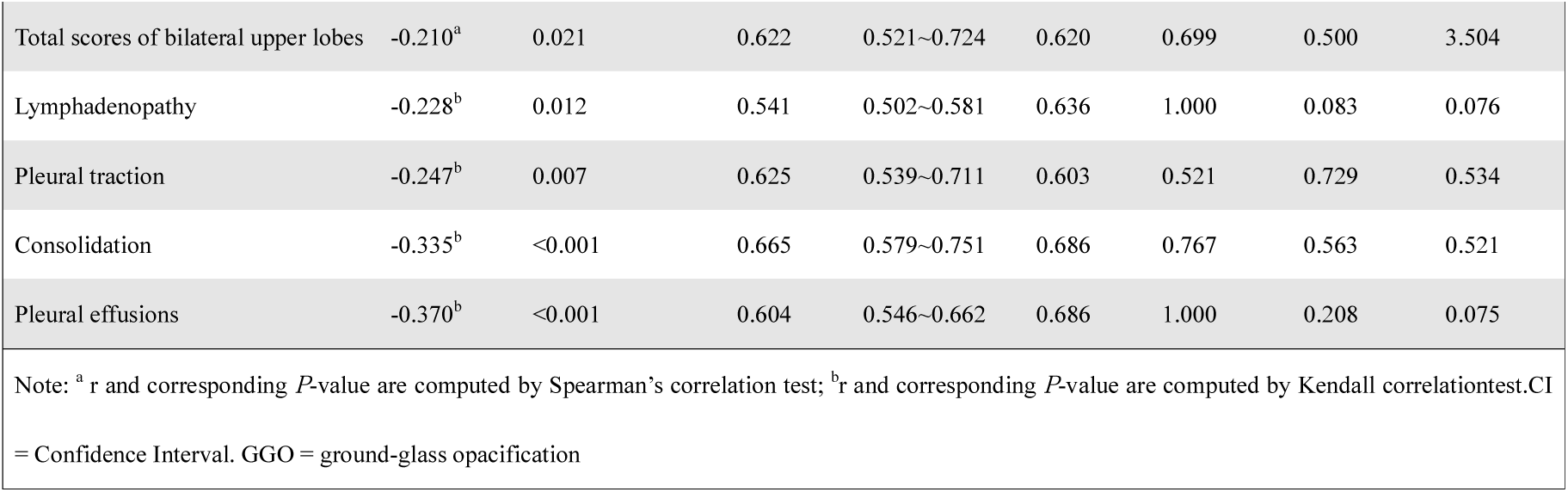
Correlation analysis and diagnostic performance of CT features in distinguishing COVID-19 from influenza pneumonia

**Figure 5.**
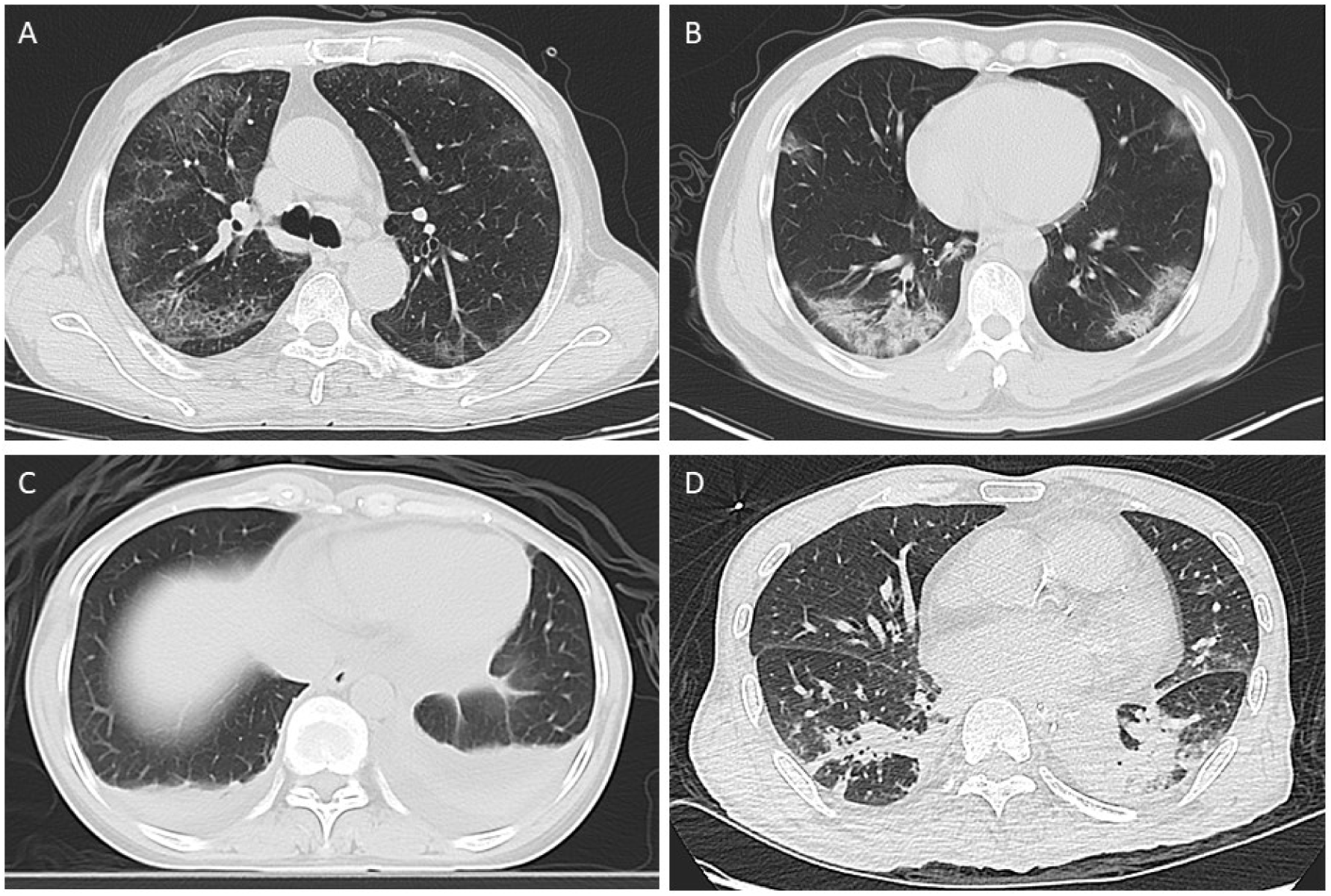
The typical CT imaging features both in COVID-19 patients (A and B) and influenza pneumonia patients (C and D). A is a 65-year old man with fever for 4 days. Axial chest CT image shows the crazy-paving pattern sign in the posterior segment of the right upper lobe, along with bilateral peripheral multi-focal ground-glass opacities (GGO). B is a 46-year old man with cough for 2 days. Axial chest CT image shows multi-focal mixed-GGO in the lower lobe of both lungs, mainly peripheral. C is a 44-year old female presenting with fever for 3 days. Axial chest CT image shows consolidations in the posterior basal segment of both lungs, along with bilateral pleural effusions. D is a 60-year old man with cough for 2 days. Axial chest CT image shows local consolidations in the dorsal segment of both lower lobes.

## Discussion

In this study, we compared CT imaging and clinical manifestations between COVID-19 and influenza pneumonia and identified the most valuable features for differential diagnosis. Among a total of 62 features, 20 imaging features and 6 clinical features were found to be significantly different. Correlation analysis showed that the WBC count had the highest correlation (r = −0.526, *P* < 0.001), with a threshold of 6.435 × 10^9^/L, followed by neutrophil count (r = −0.500, *P* < 0.001). Four CT imaging features were identified as the most significant for differential diagnosis in the early stage of both diseases, including crazy paving pattern, mixed ground-glass opacity in peripheral area, pleural effusions, and consolidation.

The GGO in the periphery has become a recognized indicator of COVID-19 in the early stage (5, 16, 17). In line with previous studies, we found that in the early stage of COVID-19, about 78% of patients had mixed GGO. However, this feature only ranked the third among the 26 extracted features for distinguishing COVID-19 from influenza. Crazy paving pattern, which also been reported in previous studies (3, 18, 19), was considered to be the most powerful feature for the differential diagnosis. These two features were also reported in other coronavirus diseases, such as severe acute respiratory syndrome (SARS) and middle east respiratory syndrome (MERS) (20, 21). The pathology of COVID-19 was confirmed to greatly resemble those of SARS and MERS (22, 23). Tian et al. reported that the lungs of COVID-19 patients exhibited edema, proteinaceous exudate, focal reactive hyperplasia of pneumocytes with patchy inflammatory cellular infiltration, and multinucleated giant cells (24), which can cause the thickening of interlobular septa, and represented as crazy paving pattern. Consistent with previous reports (25), the pleural effusions are very rare in COVID-19 patients, which ranked the second among CT imaging features for differential diagnosis.

Unlike COVID-19, influenza viruses are members of the Orthomyxoviridae family. The pathogenesis of influenza is the destruction of airway epithelial barrier, resulting in necrotizing bronchitis and diffuse alveolar damage (26). The common imaging findings of influenza are consolidation and Bronchial wall thickening (26, 27). Consistent with previous reports, we found that over 56 percent of influenza patients had positive consolidation, while the positive rate is only 23 percent for COVID-19 patients in the early stage. The positive rate is significantly different with P-value less than 0.001. However, Bernheim et al. found that in a longer time after onset, more consolidation was presented in COVID-19 patients (28), which was also confirmed by Shi et al. (5). Therefore, in the follow up of the disease, the difference of this feature between the two diseases may be weakened. The bronchial wall thickening was proved to be not significantly different between influenza and COVID-19 pneumonia (*p* = 0.715), which indicated that both diseases could affect airway walls.

Recently, RT-PCR and serological antibody tests are widely adopted for COVID-19 diagnosis. However, false-negative cases using RT-PCR have been reported in several studies (29-31). Serum antibody test was shown to have good performance for the diagnosis of COVID-19, with sensitivity of 88.66% and specificity of 90.63% (32). Because it likely takes the body one to three weeks to produce the antibodies, antibody test is unable to diagnose the illness in the early stage. To et al (33) found that IgG or IgM antibody increased for most patients at 10 days or later after symptom onset. Therefore, imaging and clinical findings have the advantage to reflect the disease earlier. To our best knowledge, our study is the first to evaluate the significant statistical difference of CT imaging and clinical features between COVID-19 and influenza pneumonia. It is worth noting that every individual feature has limited diagnostic efficacy and thus the combination of multiple features will be the trend of future research.

There are several limitations in this study. First, in order to evaluate the differential diagnosis in the early stage, we only compare the initial CT scanning both in COVID-19 and influenza pneumonia. Since the CT manifestations change with the course of the disease (34), our results may have a bias at different time windows. Second, there may be some inherent deviations in the multi-center retrospective design (35), since the scanning protocols are slightly diverse in different hospitals. Finally, although the preliminary results are promising, further validation on a larger and independent dataset is needed to determine the potential of these features for distinguishing COVID-19 from influenza pneumonia. After validation, further diagnostic models may be created based on these features.

In conclusion, a total of 1537 lesions and 62 features were compared between COVID-19 and influenza pneumonia patients. Twenty-six features were significantly different between the two groups. In CT imaging, the crazy paving pattern was recognized as the most powerful feature in the differential diagnosis in the early stage, with AUC of 0.687 (95% CI: 0.611∼0.764). In clinical manifestations, white blood cell count had the highest AUC of 0.811 (95% CI: 0.731∼0.890). These findings help to distinguish COVID-19 from influenza pneumonia.

## Data Availability

All data are available from the authors upon reasonable request.

## Acknowledgments

This work was supported by grants from the Natural Science Foundation of China [grant number 81471730, 31870981] to R.W.; the 2020 LKSF cross-disciplinary research grants [grant number 2020LKSFBME06]; the Natural Science Foundation of Guangdong Province [grant number 2018A030307057] to Z.D.; and the Special Project on Prevention and Control of COVID-19 for Colleges and Universities in Guangdong Province (grant number 2020KZDZX1085) to Z.D.

## Article Type

Original research

## Abbreviations

COVID-19: coronavirus disease 2019
GGO: ground-glass opacity
RT-PCR: reverse transcription polymerase chain reaction
WBC: white blood cell
CRP: C-reactive protein
AUC: area under the curve
SARS: severe acute respiratory syndrome
MERS: middle east respiratory syndrome.

